# Evaluating the impact of the increase in funding entitlement to Early Learning and Childcare from 600 to 1140 hours per year on family wellbeing in Scotland: statistical analysis plan

**DOI:** 10.1101/2025.07.14.25331275

**Authors:** Jim Lewsey, Daniel Mackay, Grant Aitken, Helen Patterson

**Author notes:** **Corresponding author:** Jim Lewsey. Authors’ contributions: JL drafted the statistical analysis plan, and JL, DM, GA, and HP critically reviewed the draft manuscript, and GA and JL prepared the final draft. GA and JL led on the design of the statistical analyses. All authors critically reviewed and approved the final version of the manuscript for publication. **Competing interest:** none declared.

## Abstract

This report provides the statistical analysis plan (SAP) for a study evaluating the impact of the increase in funding entitlement to Early Learning and Childcare from 600 to 1140 hours per year on family wellbeing in Scotland. In order to measure the outcomes for children, parents and carers, and families as a whole, this study is part of a larger outcomes-based evaluation that also includes an evaluation of the accessibility, flexibility, affordability, quality and take up of funded ELC and an assessment of the economic costs and benefits of the expansion. The expansion aims to contribute to three high-level outcomes:

- children’s development improves and the poverty-related outcomes gap narrows
- parents’ opportunities to take up or sustain work, study or training increases
- family wellbeing improves

## Background

From August 2021 onwards, the Scottish Government (SG) funded entitlement to early learning and childcare (ELC) in Scotland increased from 600 to 1140 hours per year for all three-and four-year-olds, and eligible two-year-olds (1). The main aim of this expansion of funded ELC was to achieve improved outcomes for children. The impacts on different groups of children and parents will be considered in relation to this aim, particularly those at risk of disadvantage, including levels of area deprivation, urban or rural areas, parents in different income groups, children with different ages, and children with additional support needs. In October 2022, SG published an evaluation strategy (2) that laid out plans for evaluating this increase of provision, which listed family wellbeing as one of three high-level outcomes to be evaluated.

## Aim and research questions

### Aim

To estimate the impact the expansion of funded ELC from 600 to 1140 hours per year has had on family wellbeing and how this varies by demographic group.

Research questions (RQs)

1. How do demographics and family wellbeing outcomes of eligible two-year-olds as they start ELC compare between November 2018 (entitled to 600 hours group) and November 2023 (entitled to 1140 hours group)?
2. How do demographics and family wellbeing outcomes of three-year-olds who were eligible at age 2 after 1 year of ELC compare between November 2019 (entitled to 600 hours group) and November 2024 (entitled to 1140 hours group)?
3. How do demographics and family wellbeing outcomes of three-year-olds starting ELC compare between November 2019 (entitled to 600 hours group) and November 2024 (entitled to 1140 hours group)?
4. How do demographics and family wellbeing outcomes of three-year-olds compare between those in November 2019 (entitled to 600 hours group) who had baseline data collected one year prior and those starting ELC in November 2024 (entitled to 1140 hours group)?
5. How do changes in family wellbeing outcomes between eligible two-year-olds as they start and after 1 year of ELC when they are three, compare between a cohort starting in November 2018 (entitled to 600 hours group) and a cohort starting in November 2023 (entitled to 1140 hours group)?
6. How do family wellbeing outcomes in four/five-year-olds as they leave ELC to begin school compare between May/June 2019 (entitled to 600 hours group) and May/June 2024 (entitled to 1140 hours group)?

## Methods

### Study design

Quantitative study; mixture of repeated cross-sectional and longitudinal designs.

### Data source

Scottish Study of Early Learning and Childcare (SSELC) – phases 1 to 6 (3). Figure 1 shows how the phases of SSELC relate to ELC provision and calendar time.

**Figure 1:**
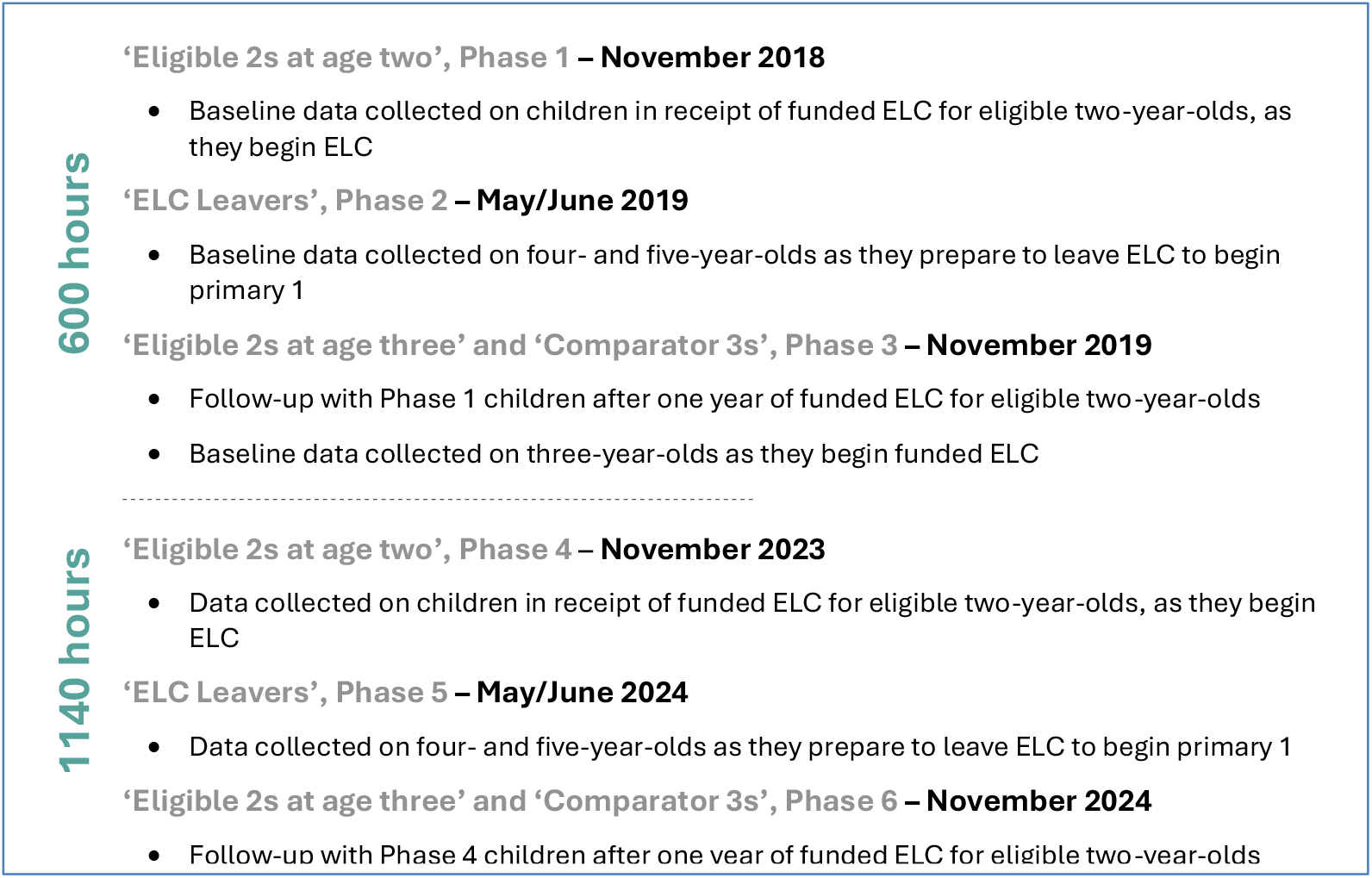
Schematic of SSELC phases and ELC provision

SSELC has several weighting strategies to address the representativeness of the cohorts. Groups that are under-represented in the achieved sample are given higher weights than those that are over-represented, with the aim of weighted data matching the population distribution by key characteristics. ELC settings in the most deprived areas for Phases 2 and 5 were deliberately oversampled (4).

Three sets of weights were produced for setting head responses, keyworker responses and parent responses. The same basic weighting approach was used for all three sets of weights, with specific modifications where required, a methodology which was consistent across all phases of SSELC. We will undertake statistical inference from both unweighted and parent weighted responses to understand the impact of different weighting schemes and ensures the validity of the results.

### Outcome measurement

Family wellbeing is a concept; a latent variable with no universally agreed definition. It is recognised as being multi-dimensional and the definition will differ depending on the context. There was no single question asked in SSELC which related directly to the concept family wellbeing.

Instead, the evaluation team at PHS identified 10 questions in the SSELC which are relevant to the concept of family wellbeing. These questions were grouped across three areas (parent health and wellbeing, home environment and parent-child relationship) and are detailed in Table 1.

**Table 1:** Questions in SSELC relating to family wellbeing.

| Parent health and wellbeing | Home environment | Parent-child relationship |
| --- | --- | --- |
| **Question (Q) 1.** How is your health in general? (Very good, Good, Fair, Poor, Very poor)<br>**Q2.** Do you have a physical or mental health condition or illness lasting or expected to last 12 months or more? (Yes, No)<br>**Q3.** Does this condition or illness limit your activities in any way? (Yes a lot, Yes a little, No)<br>**Q4.** SWEMWBS – mental wellbeing scale (7.00 – 35.00)<br>**Q5.** Life satisfaction scale (0 – 10)<br>**Q6.** How do you feel about the amount of support/help with childcare you get from family or friends living outside your household (I don't need any support, I get enough support, I don't get enough support, I don't get any support at all) | **Q7.** CHAOS (Confusion, Hubbub and Order Scale) (4 – 20)<br>**Q8.** Home learning environment scale (0 – 28) | **Q9.** Coping as a parent (Coping well as a parent most or all the time, Coping well as a parent less often)<br>**Q10.** Parent-child warmth scale (7 – 35) |

**Table 2:** Information on scales used in family wellbeing outcomes.

| Scale | Range | Interpretation | Reference |
| --- | --- | --- | --- |
| Shortform Warwick-Edinburgh Mental Wellbeing Scale (SWEMWBS) | Scores range between 7 and 35<br>Comprises of responses to 7 items, with a score of 1 for “none of the time” and 5 for “all of the time” | High wellbeing is defined as the top 15% of scores, in the range from 27.5 to 35.0, and low wellbeing as the bottom 15%, in the range from 7.0 to 19.5. | WEMWBS website, https://warwick.ac.uk/services/innovations/wemwbs, accessed 22 May 2025 |
| Home learning environment scale (HLE) | Scores range between 0 to 28<br>Score are derived by summing the number of days on which each of four activities between the child and parent occurred in the last week | Lower scores indicate less activities in the last week.<br>0 to 16 – bottom quartile<br>17 to 21 – second quartile<br>22 to 24 – third quartile<br>25 to 28 – top quartile<br>Quartiles above were from data from Phase 2 of a study (see Niklas et al 2016). | Niklas, F., Nguyen, C., Cloney, D.S. *et al.* Self-report measures of the home learning environment in large scale research: Measurement properties and associations with key developmental outcomes. *Learning Environ Res* **19**, 181–202 (2016).<br>https://doi.org/10.1007/s10984-016-9206-9 |
| Parent-child warmth scale | Scores range between 7 and 35<br>Comprises the seven items that form the warmth dimension of the short form of the Mothers’ Object Relations Scale (MORS-SF).<br>Each item is scored from 1 to 5, with 1 being “None of the time” and 5 “All of the time”. | Low scores indicate less warmth.<br>7-31 – bottom tertile<br>32 to 34 – middle tertile<br>35 – top tertile<br>Skewed data – based on responses to Phase 1 of a study (see MORS website) | Parent-Child Warmth Scale, derived from the warmth dimension of the Mothers’ Object Relations Scale (MORS-SF), see: https://www.morscales.org/mors-tools/, accessed 22 May 2025 |
| Confusion, Hubbub and Order Scale (CHAOS) | Scores range between 4 and 20<br>Comprises responses (between 1 and 5) to each of 4 statements used to measure calmness and order within the household. | Lower values indicate calmer households and higher values indicate a greater degree of disorganisation / lack of routine | Adam P. Matheny, Theodore D. Wachs, Jennifer L. Ludwig, Kay Phillips,<br>Bringing order out of chaos: Psychometric characteristics of the confusion, hubbub, and order scale, Journal of Applied Developmental Psychology, Volume 16, Issue 3, 1995, Pages 429-444, ISSN 0193-3973, [https://doi.org/10.1016/0193-3973(95)90028-4](https://doi.org/10.1016/0193-3973%2895%2990028-4). |
| Life satisfaction scale | In the SSELc study respondents are asked how satisfied they are with their life nowadays on a scale of 0 to 10. | Higher score, higher life satisfaction | SSELc Phase 5 Report, Scottish Government |

To measure family wellbeing it was decided to use a latent class analysis approach, a statistical procedure used to identify qualitatively different subgroups, using unobserved subgroups within the data based on patterns of responses to the ten questions in Table 1.

### Statistical analyses

#### Overall approach

To measure family wellbeing it was decided to use a latent class analysis (LCA) approach using unobserved subgroups within the data based on patterns of responses to the ten questions in Table 1.

#### Data preparation

Using Stata code, we will prepare 6 analysis data sets, one for each RQ. RQs 1-4 will be answered using appropriate descriptive statistics.

#### Methodological approach

RQs 5-6 will be answered by both descriptive and inferential statistical approaches. It was decided to use LCA inferential methods. This was considered appropriate due to family wellbeing being considered a latent variable. As the responses to the 10 family wellbeing questions (see Table 1) provide a mixture of categorical and numerical variables, we decided to use a generalised structural equation modelling (GSEM) approach (5) to undertake LCA using models where outcome variables are assumed to have Normal (Q4, Q5, Q7, Q8 and Q10 in Table 1), Ordinal (Q1, Q3, Q6 and Q9) and Binomial (Q2) distributions. In LCA, models are developed with different total numbers of classes. We will use a combination of statistical (AIC) and theoretical (subject expertise) criteria to determine which number of classes provides the ‘best’ solution.

For RQ5, the outcome variables in the GSEM corresponding to the 10 family wellbeing questions are constructed as a difference between responses at one year after ELC and at beginning of ELC. For the numerical variables (Q4, Q5, Q7, Q8 and Q10 in Table 1) the differences are also treated as numerical variables. For the categorical variables (Q1, Q2, Q3, Q6 and Q9 in Table 1) the differences are constructed as ordinal variables with three categories, namely ‘worse’, ‘same’ and ‘improved’. For example, if an individual has a response to Q1 of ‘Fair’ at beginning of ELC and ‘Very good’ after one year of ELC the difference would be measured as ‘improved’.

#### Representation of intervention and potential confounding in models

An intervention covariate will be used in the models to estimate the effect of extra ELC provision (0 = 600 hours, 1 = 1140 hours). The research team will identify a list of potential confounders, measured in SSELC, to adjust for in the statistical models. To potentially confound, the variable must be associated with the intervention covariate and associated with one or more of the family wellbeing outcomes. If we find evidence of collinearity between confounding variables, we will decide within the research team which of the pair of collinear variables to retain in the modelling based on criteria such as level of missing data, likely measurement error, etc.

#### Missing data

One type of missing data stems from whether the parent questionnaire was completed or not. The analyses are based on data sets where the answer to this question was ‘yes’.

Demographics between those who answered ‘yes’ and ‘no’ will be compared to see if there are any systematic differences. Across most outcome measures and potential confounding variables the level of missing data was small and therefore the analyses will proceed using a complete cases approach (6).

#### Expressing uncertainty in the results

Using GSEM there will be intervention effect sizes presented with corresponding 95% confidence intervals for the 10 outcomes (see Table 1). For the best LCA solution, we will present how the intervention groups are associated with the probabilities of class membership. To propagate uncertainty in this inference we will use a non-parametric bootstrap (7).

#### Data presentation

First, we will present the sample characteristics and responses to the indicator variables, highlighting any differences between SSELC data collection phases. Second, for each RQ, we will identify latent profiles and display rationale for choice of models based on multiple fit statistics and theoretical interpretability. Third, for each RQ, we will compare allocation of class probabilities against each of the questions in Table 1 plus additional variables of equivalised household income quintiles, degree level education, lone parent household, if ‘on schedule’ by Ages and Stages Questionnaire (ASQ) domain, and if ‘close to average’ Strengths and Difficulties Questionnaire (SDQ) domain. Finally, we will display relative risks of being in each class after exposure to increased hours.

## Data Availability

An application for data access would need to be made with the Scottish Study for Early Learning and Childcare (SSELC)

## Notes

**Funding:** This work is supported by funding from PHS as part of a service level agreement between PHS and the University of Glasgow (‘The Provision of Specialist Statistical Advice and Support Services’), 2025.

### Competing Interest Statement

The authors have declared no competing interest.

### Funding Statement

This study did not receive any funding.

## References

1. Government S. Early education and care - Funded early learning and childcare [Available from: https://www.gov.scot/policies/early-education-and-care/early-learning-and-childcare/.

2. Government S. Early learning and childcare expansion programme: evaluation strategy. 2022.

3. Government S. Collection - Early learning and childcare expansion evaluation [Available from: https://www.gov.scot/collections/early-learning-and-childcare-expansion-valuation/#scottishstudyofearlylearningandchildcare.

4. Hinchcliffe, S, et all (2021). Scottish Study of Early Learning and Childcare: Three-year-olds (Phase 3) Report. Scottish Centre for Social Research

5. Acock, A. C. (2013). Discovering Structural Equation Modeling Using Stata (Revised ed.). College Station, TX: Stata Press.

6. Mukaka, M, et all (2016). Is using multiple imputation better than complete case analysis for estimating a prevalence (risk) difference in randomized controlled trials when binary outcome observations are missing? Trials.

7. Caers, J, et all (1998). Nonparametric tail estimation using a double bootstrap method. Computational Statistics & Data Analysis, Volume 29, Issue 2.

